# Integrating Genetic Ancestry into Clinical Care: Accuracy, Utility, and Stakeholder Views

**DOI:** 10.1101/2024.05.01.24306719

**Authors:** Jessica Prettyman, Thomas J Hoffmann, Sawona Biswas, Aleksandar Rajkovic

## Abstract

**PURPOSE:** This study aimed to evaluate the concordance of genetic ancestry reports from different providers, assess the accuracy of genetic ancestry compared to self-identified race and ethnicity (SIRE), and explore patient and provider perspectives on the potential utility and integration of genetic ancestry data into the electronic health record (EHR).

**METHODS:** Genetic ancestry results from two commercial providers and two 3rd-party analyses were compared for concordance using data from 451 participants in the UCSF 3D Health Study. Genetic ancestry was also compared to SIRE. Surveys were administered to gather perspectives on genetic ancestry testing, its accuracy, and potential integration into the EHR.

**RESULTS:** The overall mean concordance between the two commercial providers was 58.41%. Ancestry from one provider had the highest concordance with SIRE, ranging from 80.05% to 94.78% across different thresholds. The majority of participants and providers were neutral regarding the integration of genetic ancestry into the EHR.

**CONCLUSION:** Significant discordance exists between genetic ancestry reports from different providers, highlighting the need for standardization in the calculation of genetic ancestry. While participants and providers acknowledge the potential utility of genetic ancestry in personalized medicine, concerns regarding data privacy, accuracy, and the potential for discrimination must be addressed before integration into the EHR.

## INTRODUCTION

The use of race and genetic ancestry in clinical care has been a topic of intense debate in recent years.^1–10^ Race is a social construct that does not directly correspond to biology or genetics, providing limited information about an individual’s potential genetic predispositions.^6^ In contrast, genetic ancestry utilizes genetic information to identify populations sharing common genetic characteristics, which can play a role in understanding an individual’s risk for various health conditions.^6,8,10–12^

Currently, self-identified race and ethnicity (SIRE) are the most commonly used proxies for genetic ancestry in healthcare. However, genetic testing providers develop their ancestry tests independently, leading to potential inconsistencies among reports from different companies.^8,13,14^ These discrepancies can lead to confusion among healthcare providers and consumers regarding the accuracy and reliability of these tests. Limited research has investigated the concordance of genetic ancestry tests, with existing studies having limitations in sample size, diversity, and the number of commercial providers evaluated.^13,15–18^ There is a need for further research to better characterize the discordances between genetic testing providers and SIRE.

Establishing the clinical utility of genetic ancestry information is crucial for its potential integration into the electronic health record (EHR). It is essential to understand if providers would find this information valuable, if they would use it in clinical decision-making, and if they trust the validity of the data. Studies exploring provider perspectives on genetic ancestry testing are limited, with no research examining their views on its potential utility in the EHR.^19–22^ Literature on patient perspectives has found mixed opinions on genetic ancestry testing and its impact on personal identity.^23–26^

The increasing availability of genetic testing is driving a growing interest in personalized medicine.^27^ Genetic ancestry information can offer insights into an individual’s personalized risk for developing monogenic, polygenic, and multifactorial conditions. However, the validity and clinical impact of these tests are still under investigation, primarily due to the limited availability of diverse genetic data. As the use of these tests expands and their utility improves, genetic counselors will play a crucial role in interpreting and communicating these results to patients and non-genetic providers.

This study aimed to explore the utility and implications of genetic ancestry data through the perspectives of participants in the UCSF 3D Health Study and UCSF providers. The 3D Health Study, launched in 2019, investigates the predictive value of genome sequencing (GS) in a healthy adult population. Understanding the current limitations of genetic ancestry reports and the perspectives on their clinical and personal utility is essential for evaluating existing attitudes among patients and providers and informing the responsible integration of this information into healthcare.

## MATERIALS AND METHODS

This project consisted of two separate analyses, a statistical analysis of genetic ancestry data gathered from two commercial genetic ancestry providers and an analysis of provider and participant surveys.

### Institutional Review Board Approval

This project was performed with approval from the University of California, San Francisco (UCSF) Institutional Review Board (IRB). Modifications were made to the existing 3D Health Study protocol to distribute the additional surveys and perform the secondary concordance analysis. The 3D Health Study at the University of California, San Francisco (UCSF) was launched in 2019 as a pilot project to investigate the predictive value of genome sequencing (GS) in a healthy adult population. With an enrollment of 535 participants, the study aims to establish institutional best practices for obtaining, storing, and communicating genomic information. In addition to understanding the utility of WGS for healthy adults, the study focuses on determining the types of genomic results participants want to receive and how to better engage clinicians, such as primary care providers, and participants from diverse backgrounds.

### Genetic Ancestry Concordance Analysis

The data produced from the genetic ancestry analysis of the two genetic testing providers was analyzed for concordance in reports. There was a total of 451 participants with genetic ancestry results from both commercial providers which were analyzed. There were two separate concordance analyses performed on the genetic ancestry and Apex-reported race data: a comparison between Apex-reported data and genetic ancestry and a comparison of genetic ancestry calculations.

The methods of the first analysis comparing the two genetic testing providers were developed from a prior study performed on genetic ancestry data from direct-to-consumer companies.^13^ We looked for the overall concordance in geographic ancestries reported by the two genetic testing providers to better quantify the variation of genetic ancestry results. The companies reported genetic ancestry at different geographical levels. Ancestry 1 determines genetic ancestry at a more specific geographic location. Ancestry 2 reports genetic ancestry for five geographic regions; African, Admixed American, East Asian, European, and South Asian. For the analysis, we grouped the reported ancestries for Ancestry 1 into the five categories determined by Ancestry 2 to run the comparison (Supplemental Table 1). Participants were reported to have genetic ancestry from five geographic regions including African (AFR), Admixed American (AMR), East Asian (EAS), European (EUR), and South Asian (SAS). The concordance analysis aimed to determine the percent agreement between the two genetic ancestry providers for each participant and then determine the overall agreement of the two providers’ reports. The analysis performed on the data is below.

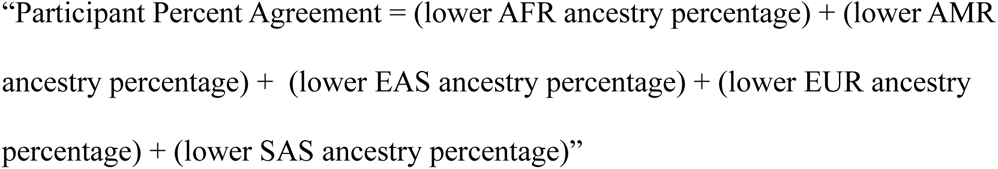

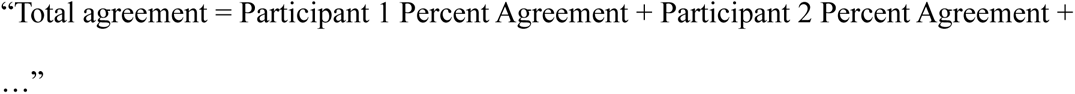

The agreements between the providers as well as between different ancestry groups were then compared. A Kruskal-Wallis test was used to determine if there were statistically significant differences between genetic ancestry groups. Pair-wise comparisons were run using a Wilcox Rank Sum test to identify statistically significant differences when comparing the mean concordance of ancestry groups.

The second analysis compared the genetic ancestry data to Apex-reported ancestry. The Apex- reported ancestry was gathered for each participant in the 3D Health study. Apex includes seven options for race/ethnicity including Hispanic or Latino, Black or African American,

White/Caucasian, Asian, Native Hawaiian or Other Pacific Islander, American Indian or Alaska Native, and Mixed. More than one race/ethnicity can be selected within Apex. For the analysis, these race/ethnicity options were combined into 5 categories that reflected the genetic ancestry results produced by each genetic testing provider. These categories were Asian (AS), African (AFR), Admixed American (AMR), European (EUR), and no data (ND). Asian included all those whose Apex-reported race was Asian, or Native Hawaiian, or Other Pacific Islander. African included those that were reported Black or African American. Admixed Americans included those whose race/ethnicity was Hispanic or Latino, or American Indian or Alaska Native. Those who selected mixed were placed into no data because their race could not be accurately evaluated. Individuals with only one race/ethnicity selected would be identified to be 100% of that race/ethnicity. Individuals with more than one race/ethnicity would have their Apex-reported ancestries evenly distributed into every race/ethnicity selected. An example of this would be if an individual reported both Asian and European races, for the concordance analysis they would be 50% Asian and 50% European. Although this method does not likely represent their true ancestry distribution, this distribution represents how healthcare providers or clinical care algorithms may utilize the Apex-reported race in care. The genetic ancestry data was then compared to the Apex- reported race of all the participants in the UCSF 3D Health Study. The methods of the analysis were based on a prior study that analyzed the concordance between Apex-reported race and genetic ancestry for those of European or African ancestry.^15^ Specifically, we calculated whether provider genetic ancestry was concordant or not for each of the three thresholds (T) of concordance, T: 50%, 75%, and 90%. For individuals reporting one race/ethnicity, genetic ancestry was considered concordant if it was ≥T% that race/ethnicity. For the individuals who reported multiple races/ethnicities, individuals were considered concordant if both corresponding genetic ancestries ≥(T/2)%. The concordances at each threshold were then compared for each ancestry provider using a matrix of McNemar’s test to identify if there was a statistically significant difference between the results.

### 3D Health Participant Survey

The 3D Health Participant Survey aimed to gather patients’ perspectives on genetic ancestry testing, its perceived benefits, drawbacks, and impact on identity. The questionnaire was sent to 527 active 3D Health participants who had not withdrawn, been lost to follow-up, or died since enrolling. An initial email asked about their interest in receiving genetic ancestry results and informed them about the additional questionnaire. Interested participants received their results and the questionnaire, to be completed after viewing the results. The survey, adapted from Rubanovich et al., included Likert-scale, yes/no, and short-response questions.^28^ It focused on the impact of genetic ancestry on identity, accuracy of results, integration into the EHR, and perceived benefits/concerns (Supplemental Materials). Quantitative and qualitative metrics were used to analyze responses. Descriptive statistics summarized demographics, perspectives on EHR integration, utility, impact on identity, and common concerns/benefits. Fisher’s exact tests determined statistically significant differences between racial/ethnic groups.

### UCSF Provider Survey

The UCSF Provider Survey explored provider perspectives on genetic ancestry testing, clinical utility, benefits, and drawbacks. A total of 128 UCSF providers in internal medicine, family medicine, genetics, genetic counseling, and medical genetics were contacted to participate. The survey, partially adapted from Nelson et al., included Likert-scale, yes/no, and short response questions.^22^ It focused on race and ancestry’s influence on medicine, EHR integration, clinical use of genetic ancestry, and concerns about unintended uses (Supplemental Materials).

Quantitative and qualitative metrics summarized demographics, perspectives on EHR integration, utility, and common concerns/benefits. Fisher’s exact tests identified statistically significant differences between provider groups.

## RESULTS

Data for this study were collected from the UCSF 3D Health Study, which aims to investigate the predictive value of whole genome sequencing in a healthy adult population and to establish best practices for obtaining, storing, and communicating genomic information.

### Respondent Demographics

#### 3D Health Participant Survey

The majority of the respondents to the study identified as women (60.8%), 60 or above (49.4%), and white (74.7%). Participants identifying as Asian represented 20.5% of the respondents, while 5.4% identified as Hispanic/Latino, 2.4% as Black/African American, and 0.6% as American Indian/Alaska Native (Supplemental Table 2).

### UCSF Provider Survey

A total of 31 UCSF providers responded to the survey, with the majority identifying as women (80.6%), white (67.7%), 30-45 years old (58.1%), and genetic counselors (45.2%) (Supplemental Table 4). The majority of participants had never had genetic ancestry testing (83.9%). Genetic counselors were the majority of the respondents (45.2%), followed by internal medicine providers (38.7%), family medicine providers (6.5%), and geneticists (3.2%) (Supplemental Table 3).

### Concordance Analysis

The concordance analysis utilized genetic data from 451 participants in the UCSF 3D Health Study to compare genetic ancestry results from two commercial providers and a 3rd-party analysis. The analysis aimed to determine the concordance between the different genetic ancestry calculations and to compare genetic ancestry results with self-reported race/ethnicity data.

### Concordance between Genetic Ancestry Providers

The concordance analysis compared the genetic ancestry results from two different commercial genetic testing providers, referred to as Ancestry 1 and Ancestry 2, for 451 samples in the UCSF 3D Health Study. The overall mean concordance between Ancestry 1 and Ancestry 2 was found to be 58.41% (Table 1). The concordance was then calculated for each genetic ancestry category: European, Asian, Admixed American, and African. Among these categories, those with European genetic ancestry were found to have the lowest mean concordance of 54.73% between Ancestry 1 and Ancestry 2. Asian ancestry had a mean concordance of 73.07%, and Admixed American ancestry had a mean concordance of 57.55%. African ancestry was found to have the highest mean concordance of 76.21% between the two genetic testing providers. A significant difference was found when comparing the overall mean concordance by ancestry (p-value = 3.2e-20). The pair-wise comparison found statistically significant differences between all of the ancestry groups except when Asian and African groups were compared (Table 1).

**Table 1.**
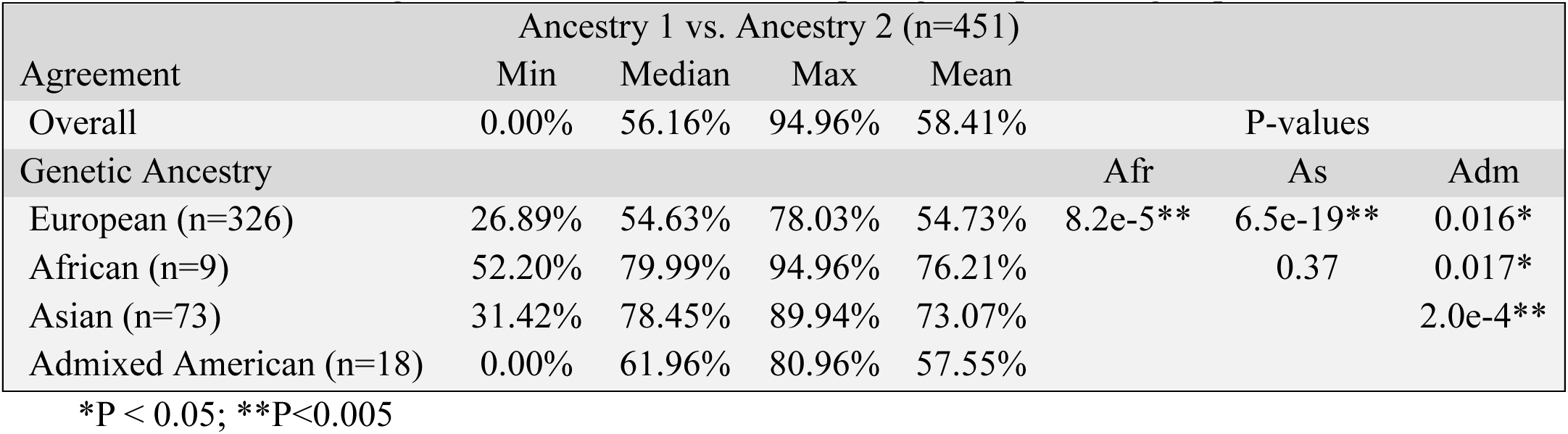
Concordance between genetic ancestry calculations. Genetic ancestry results from two different commercial genetic testing providers were compared for 451 samples in the UCSF 3D Health Study. The percent of overlapping ancestry determined the agreement from each ancestry location (European, African (Afr), Asian (As), and Admixed American (Amr)). P-values were determined using a Wilcox Rank Sum test comparing each pairwise group.

| Agreement | Ancestry 1 vs. Ancestry 2 (n=451) |  |  |  | P-values |  |  |
| --- | --- | --- | --- | --- | --- | --- | --- |
|  | Min | Median | Max | Mean |  |  |  |
| Overall | 0.00% | 56.16% | 94.96% | 58.41% |  |  |  |
| Genetic Ancestry |  |  |  |  | Afr | As | Adm |
| European (n=326) | 26.89% | 54.63% | 78.03% | 54.73% | 8.2e-5** | 6.5e-19** | 0.016* |
| African (n=9) | 52.20% | 79.99% | 94.96% | 76.21% |  | 0.37 | 0.017* |
| Asian (n=73) | 31.42% | 78.45% | 89.94% | 73.07% |  |  | 2.0e-4** |
| Admixed American (n=18) | 0.00% | 61.96% | 80.96% | 57.55% |  |  |  |

\*P < 0.05; \*\*P<0.005

### Concordance of Apex-reported Race and Genetic Ancestry

In this analysis, the genetic ancestry results from four different sources (two commercial providers, Ancestry 1 and Ancestry 2, and two supplemental ancestry calculations from a 3rd- party software, Ancestry 3 and Ancestry 4) were compared to the Apex-reported race/ethnicity for samples from the UCSF 3D Health study. Apex-reported race/ethnicity refers to the self-reported race/ethnicity information collected from participants using the UCSF Apex system.

Ancestry 1 was found to have the lowest concordance with Apex-reported race. For Ancestry 1, 69.63% of samples had at least a 50% concordance, 15.65% of samples had at least a 75% concordance, and 2.57% of samples had at least a 90% concordance with Apex-reported race (Table 2).

**Table 2.** Concordance between genetic ancestry results and apex-reported race. Genetic ancestry results from two commercial providers and two supplemental ancestry calculations from a 3^rd^-party software were compared to Apex-reported race/ethnicity for samples from the UCSF 3D Health study.

| Genetic Ancestry Calculation | Percent Concordance of Apex-Reported Race/Ethnicity by Threshold |  |  |
| --- | --- | --- | --- |
|  | 50% | 75% | 90% |
| Ancestry 1 | 69.63%<br>(298/428) | 15.65%<br>(67/428) | 2.57%<br>(11/428) |
| Ancestry 2 | 94.78%<br>(418/441) | 90.70%<br>(400/441) | 80.05%<br>(353/441) |
| Ancestry 3 | 93.42%<br>(412/441) | 90.48%<br>(399/441) | 87.98%<br>(388/441) |
| Ancestry 4 | 93.65%<br>(413/441) | 91.38%<br>(403/441) | 89.34%<br>(394/441) |

Ancestry 2 had a higher concordance with Apex-reported race compared to Ancestry 1. For Ancestry 2, 94.78% of samples had at least a 50% concordance, 90.70% of samples had at least a 75% concordance, and 80.05% of samples had at least a 90% concordance with Apex-reported race (Table 2).

Ancestry 3 and Ancestry 4 had the highest concordance with Apex-reported race among the four genetic ancestry sources. For Ancestry 3, 87.98% of samples were at least 90% concordant with Apex-reported race, while for Ancestry 4, 89.34% of samples were at least 90% concordant with Apex-reported race (Table 2).

Pair-wise comparisons of the ancestry groups found a statistically significant difference in the concordance between Ancestry 1 and every other ancestry at the 50% threshold (p-value = 5.0e- 24; 1.4e-21; 1.5e-22) and 75% threshold (p-value = 1.7e-70; 6.4e-71; 1.4e-71) (Supplemental Table 5). There is a significant difference between all of the ancestries at the 90% threshold (Supplemental Table 5).

### Participant Survey Responses

#### Impact of Genetic Ancestry on Identity

Participants were asked about their reactions to their genetic ancestry results, specifically whether the results were surprising or unexpected. The responses were evenly distributed, with 31.9% answering "Yes," 31.3% answering "No," and 35.5% answering "Somewhat/Maybe." When the responses were stratified by self-reported race/ethnicity, the majority of those identifying as Asian (51.7%) responded "No," indicating that their genetic ancestry results were not surprising or unexpected. In contrast, the majority of those identifying as African (100%), Admixed American (50%), and Other (61.5%) selected that their genetic ancestry results were somewhat or maybe surprising or unexpected. The differences in responses across self-reported race/ethnicity groups were statistically significant (p-value = 0.00653) (Table 3).

**Table 3.**
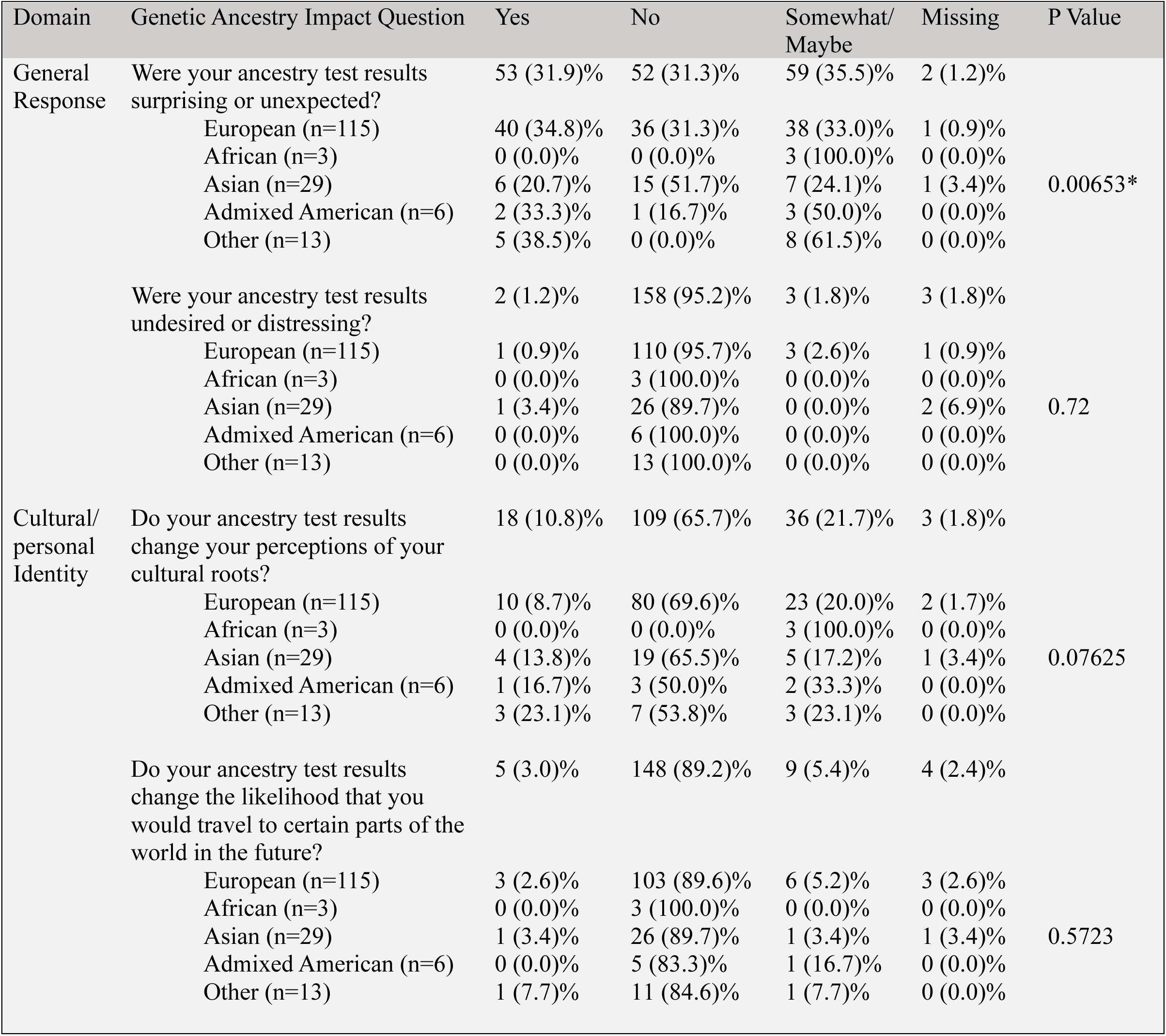
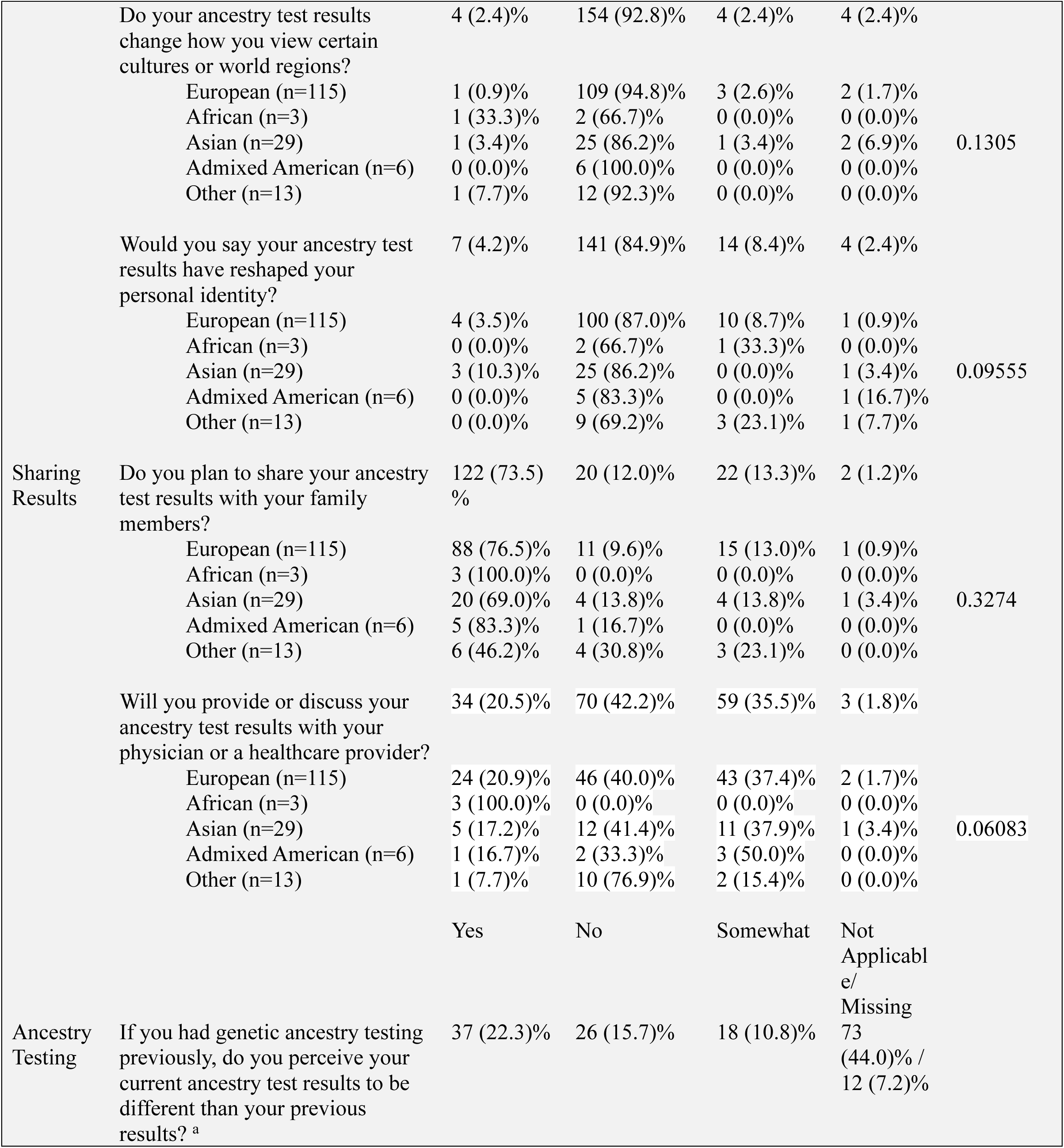
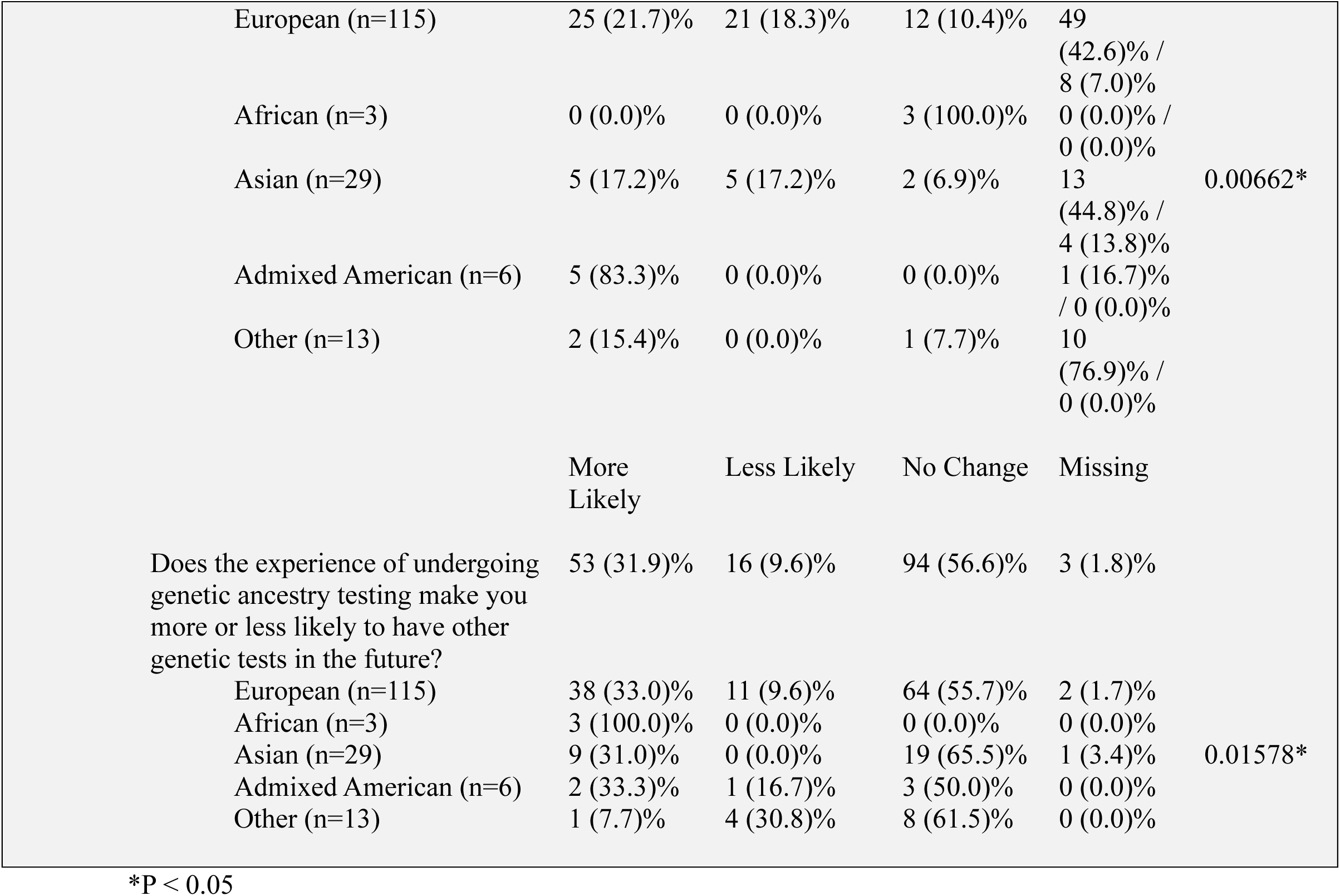
3D Health Study Participant responses to questions regarding the impact of their genetic ancestry results on identity. Genetic ancestry impact questions based on Rubanovich et al. 2021. Responses were totaled for the overall responses from all participants and then calculated based on the participant’s self-identified race/ethnicity. Participants who selected more than one race/ethnicity that did not fit into one of the general ancestry categories were placed into other. A Fisher’s Exact test was used to determine if there was a statistically significant difference in responses between ancestry groups.

Participants were also asked if their genetic ancestry reports were undesired/distressing, changed their perceptions of cultural identity, or reshaped their identity. The majority of respondents selected "No" for all three questions (95.7%, 65.7%, and 84.9%, respectively).

For respondents who had undergone genetic ancestry testing prior to the study, there was a mixed response regarding whether they perceived their current genetic ancestry results as different from their previous results.

Participants were asked if undergoing genetic ancestry testing changed the likelihood of them having other genetic tests in the future. The majority of respondents (56.6%) felt no change in the likelihood, but there was a statistically significant difference in responses across SIRE groups (p-value = 0.01578). The majority of European (55.7%), Asian (65.5%), Admixed American (50%), and Other (61.5%) respondents felt there was no change, while all respondents who identified as African (100%) felt they were more likely to have genetic ancestry testing in the future (Table 3).

### Perceived Accuracy and Utility of Genetic Ancestry

Participants were asked about their perceptions of the accuracy and utility of genetic ancestry reports. Over half of the 3D Health participants somewhat (44.6%) or strongly (6.6%) agreed that genetic ancestry reports are accurate (Table 4). When asked if genetic ancestry reports will allow for more personalized care, the majority of respondents selected neutral (30.1%), somewhat agree (33.7%), or strongly agree (16.3%).

**Table 4.**
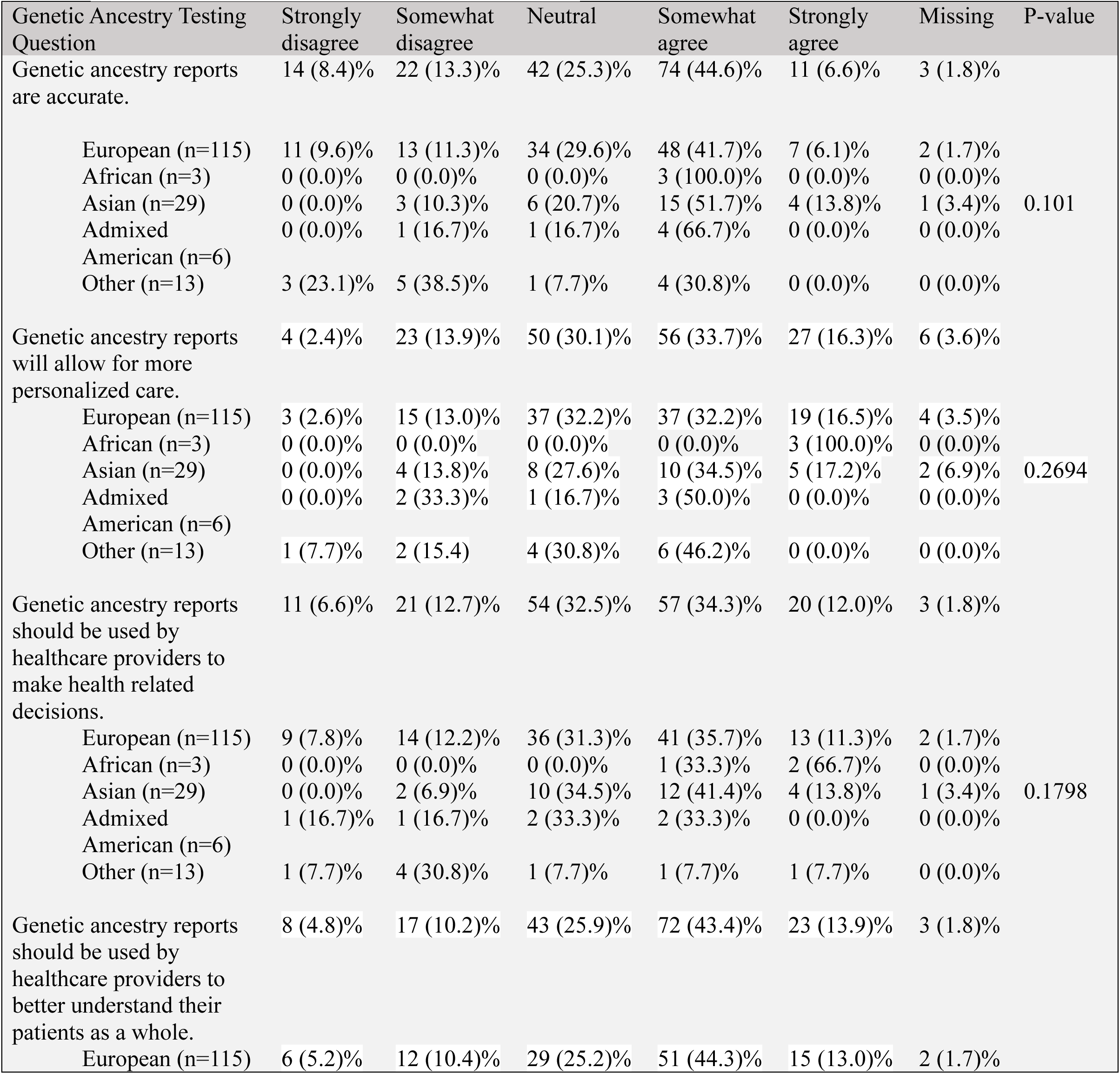
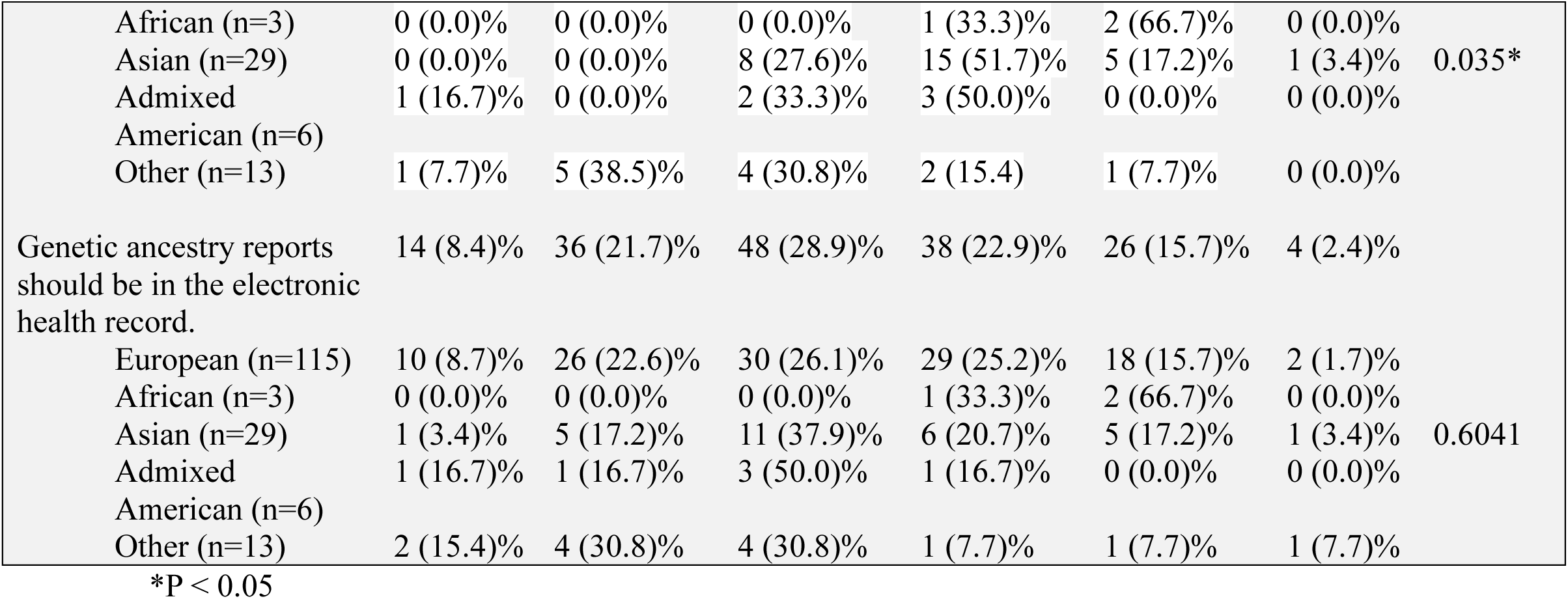
Participant responses to questions regarding genetic ancestry testing accuracy and utility. Responses were totaled for the overall responses from all participants and then calculated based on the participant’s self-identified race/ethnicity. Participants who selected more than one race/ethnicity that did not fit into one of the general ancestry categories were placed into other. A Fisher’s Exact test was used to determine if there was a statistically significant difference in responses between ancestry groups.

| Genetic Ancestry Testing Question | Strongly disagree | Somewhat disagree | Neutral | Somewhat agree | Strongly agree | Missing | P-value |
| --- | --- | --- | --- | --- | --- | --- | --- |
| Genetic ancestry reports are accurate. | 14 (8.4)% | 22 (13.3)% | 42 (25.3)% | 74 (44.6)% | 11 (6.6)% | 3 (1.8)% |  |
| European (n=115) | 11 (9.6)% | 13 (11.3)% | 34 (29.6)% | 48 (41.7)% | 7 (6.1)% | 2 (1.7)% | 0.101 |
| African (n=3) | 0 (0.0)% | 0 (0.0)% | 0 (0.0)% | 3 (100.0)% | 0 (0.0)% | 0 (0.0)% |  |
| Asian (n=29) | 0 (0.0)% | 3 (10.3)% | 6 (20.7)% | 15 (51.7)% | 4 (13.8)% | 1 (3.4)% |  |
| Admixed | 0 (0.0)% | 1 (16.7)% | 1 (16.7)% | 4 (66.7)% | 0 (0.0)% | 0 (0.0)% |  |
| American (n=6) |  |  |  |  |  |  |  |
| Other (n=13) | 3 (23.1)% | 5 (38.5)% | 1 (7.7)% | 4 (30.8)% | 0 (0.0)% | 0 (0.0)% |  |
| Genetic ancestry reports will allow for more personalized care. | 4 (2.4)% | 23 (13.9)% | 50 (30.1)% | 56 (33.7)% | 27 (16.3)% | 6 (3.6)% |  |
| European (n=115) | 3 (2.6)% | 15 (13.0)% | 37 (32.2)% | 37 (32.2)% | 19 (16.5)% | 4 (3.5)% | 0.2694 |
| African (n=3) | 0 (0.0)% | 0 (0.0)% | 0 (0.0)% | 0 (0.0)% | 3 (100.0)% | 0 (0.0)% |  |
| Asian (n=29) | 0 (0.0)% | 4 (13.8)% | 8 (27.6)% | 10 (34.5)% | 5 (17.2)% | 2 (6.9)% |  |
| Admixed | 0 (0.0)% | 2 (33.3)% | 1 (16.7)% | 3 (50.0)% | 0 (0.0)% | 0 (0.0)% |  |
| American (n=6) |  |  |  |  |  |  |  |
| Other (n=13) | 1 (7.7)% | 2 (15.4) | 4 (30.8)% | 6 (46.2)% | 0 (0.0)% | 0 (0.0)% |  |
| Genetic ancestry reports should be used by healthcare providers to make health related decisions. | 11 (6.6)% | 21 (12.7)% | 54 (32.5)% | 57 (34.3)% | 20 (12.0)% | 3 (1.8)% |  |
| European (n=115) | 9 (7.8)% | 14 (12.2)% | 36 (31.3)% | 41 (35.7)% | 13 (11.3)% | 2 (1.7)% | 0.1798 |
| African (n=3) | 0 (0.0)% | 0 (0.0)% | 0 (0.0)% | 1 (33.3)% | 2 (66.7)% | 0 (0.0)% |  |
| Asian (n=29) | 0 (0.0)% | 2 (6.9)% | 10 (34.5)% | 12 (41.4)% | 4 (13.8)% | 1 (3.4)% |  |
| Admixed | 1 (16.7)% | 1 (16.7)% | 2 (33.3)% | 2 (33.3)% | 0 (0.0)% | 0 (0.0)% |  |
| American (n=6) |  |  |  |  |  |  |  |
| Other (n=13) | 1 (7.7)% | 4 (30.8)% | 1 (7.7)% | 1 (7.7)% | 1 (7.7)% | 0 (0.0)% |  |
| Genetic ancestry reports should be used by healthcare providers to better understand their patients as a whole. | 8 (4.8)% | 17 (10.2)% | 43 (25.9)% | 72 (43.4)% | 23 (13.9)% | 3 (1.8)% |  |
| European (n=115) | 6 (5.2)% | 12 (10.4)% | 29 (25.2)% | 51 (44.3)% | 15 (13.0)% | 2 (1.7)% |  |
| African (n=3) | 0 (0.0)% | 0 (0.0)% | 0 (0.0)% | 1 (33.3)% | 2 (66.7)% | 0 (0.0)% | 0.035* |
| Asian (n=29) | 0 (0.0)% | 0 (0.0)% | 8 (27.6)% | 15 (51.7)% | 5 (17.2)% | 1 (3.4)% |  |
| Admixed | 1 (16.7)% | 0 (0.0)% | 2 (33.3)% | 3 (50.0)% | 0 (0.0)% | 0 (0.0)% |  |
| American (n=6) |  |  |  |  |  |  |  |
| Other (n=13) | 1 (7.7)% | 5 (38.5)% | 4 (30.8)% | 2 (15.4) | 1 (7.7)% | 0 (0.0)% |  |
| Genetic ancestry reports should be in the electronic health record. | 14 (8.4)% | 36 (21.7)% | 48 (28.9)% | 38 (22.9)% | 26 (15.7)% | 4 (2.4)% | 0.6041 |
| European (n=115) | 10 (8.7)% | 26 (22.6)% | 30 (26.1)% | 29 (25.2)% | 18 (15.7)% | 2 (1.7)% |  |
| African (n=3) | 0 (0.0)% | 0 (0.0)% | 0 (0.0)% | 1 (33.3)% | 2 (66.7)% | 0 (0.0)% |  |
| Asian (n=29) | 1 (3.4)% | 5 (17.2)% | 11 (37.9)% | 6 (20.7)% | 5 (17.2)% | 1 (3.4)% |  |
| Admixed | 1 (16.7)% | 1 (16.7)% | 3 (50.0)% | 1 (16.7)% | 0 (0.0)% | 0 (0.0)% |  |
| American (n=6) |  |  |  |  |  |  |  |
| Other (n=13) | 2 (15.4)% | 4 (30.8)% | 4 (30.8)% | 1 (7.7)% | 1 (7.7)% | 1 (7.7)% |  |

\*P < 0.05

Participants were also asked if genetic ancestry reports should be used by healthcare providers to make health-related decisions and to better understand their patients as a whole. 78.8% of respondents selected that they are neutral, somewhat agree, or strongly agree that genetic ancestry reports should be used for health-related decisions, and 83.2% felt the same about providers using ancestry to better understand their patients. There was a statistically significant difference in responses across SIRE groups (P-value = 0.035) (Table 4), with a higher percentage of African-identifying respondents (66.7%) strongly agreeing that providers should use genetic ancestry to better understand their patients as a whole.

### Perspectives on Integration of Genetic Ancestry into the EHR

Participants were asked if genetic ancestry reports should be included in the electronic health record (EHR). A small majority of respondents somewhat or strongly agreed with this idea (Table 4). Participants were also asked about their concerns regarding the possible unintended outcomes of including genetic ancestry data in the EHR. The most commonly selected concerns were privacy (59.0%) and misuse of genetic data (74.1%) (Supplemental Figure 2). Other concerns reported by participants included data breaches, unintentional biases, discordance between different ancestry reports, and questionable accuracy. When asked specifically about their thoughts on including genetic ancestry reports in the EHR, the majority of respondents selected privacy concerns (58.4%) and misuse of genetic data (69.9%) (Supplemental Figure 3). Additional concerns included insurance companies using the results to determine risk for pre-existing conditions or risk factors, questionable accuracy, and data breaches.

### Genetic Discrimination

Participants were asked if they expect genetic ancestry reports to be used to discriminate between groups. The responses were almost evenly split, with 52.4% responding "Yes" and 42.2% responding "No" (Table 5). There was no statistically significant difference in the responses between different SIRE groups. All respondents identifying as African responded "No," while Admixed Americans had the highest percentage of "Yes" responses (66.7%) (Table 5).

**Table 5.** 3D Health participant responses to concerns about genetic ancestry being used to discriminate between groups. Responses were totaled for the overall responses from all participants and then calculated based on the participant’s self-identified race/ethnicity. Participants who selected more than one race/ethnicity that did not fit into one of the general ancestry categories were placed into other. A Fisher’s Exact test was used to determine if there was a statistically significant difference in responses between ancestry groups.

| Do you expect that genetic ancestry reports will be used to discriminate? | Yes | No | Missing | P-Value |
| --- | --- | --- | --- | --- |
|  | 87 (52.4)% | 70 (42.2)% | 9 (5.4)% |  |
| European (n=115) | 58 (50.4)% | 49 (42.6)% | 8 (7.0)% | 0.3863 |
| African (n=3) | 0 (0.0)% | 3 (100.0)% | 0 (0.0)% |  |
| Asian (n=29) | 17 (58.6)% | 11 (37.9)% | 1 (3.4)% |  |
| Admixed American (n=6) | 4 (66.7)% | 2 (33.3)% | 0 (0.0)% |  |
| Other (n=13) | 8 (61.5)% | 5 (38.5)% | 0 (0.0)% |  |

### Provider Survey Responses

#### Perspectives on Integration of Genetic Ancestry into the EHR

Providers were asked about their perspectives on integrating genetic ancestry information into the EHR. The majority of providers felt neutral about this integration, and there was no statistically significant difference between the responses of providers from different SIRE groups. No provider strongly agreed with the statement that genetic ancestry reports should be integrated into the EHR (Supplemental Table 6).

## DISCUSSION

Our study aimed to identify the concordance of genetic ancestry reports calculated by different providers and to explore the perspectives of 3D Health Study participants and UCSF providers on the utility and implications of genetic ancestry data.

The comparison of genetic ancestry results from two commercial providers revealed an average concordance of only 58.41% (Table 1), indicating that individuals may receive notably different results depending on the provider. This finding aligns with prior studies that have found similar discordances between genetic ancestry reports.^13^ Despite both providers using the 1,000 Genomes Project database, the discrepancies in their reports demonstrate that genetic ancestry calculations can vary significantly even when using the same reference population. These inconsistencies underscore the need for standardization in the calculation of genetic ancestry before its integration into the EHR.

Participants identifying as White had the most discordant results between the two providers, with a mean concordance of 54.73% (Table 1). Contrary to our expectations, the majority of White respondents were either neutral or agreed that genetic ancestry tests are accurate (Table 6). Both participants and providers were largely neutral or in agreement regarding the integration of genetic ancestry into the EHR (Table 6; Supplemental Table 6), possibly indicating a lack of established utility for genetic ancestry at this time.

Prior studies have found genetic ancestry to be important in interpreting polygenic risk scores for an individual’s risk of polygenic and multifactorial conditions.^6,29,30^ Certain genetic ancestries have also been shown to correlate with a higher incidence of variants of uncertain significance (VUSs) in genetic testing, likely due to the underrepresentation of minority groups in genetic studies.^31^ Awareness of the increased likelihood of VUSs based on ancestry can inform pre-test genetic counseling and help manage patient expectations regarding uncertain results.

Following the distribution of genetic ancestry reports and the survey, we received numerous inquiries concerning the accuracy of the data and whether the correct reports were provided. Previous studies have found that discrepancies between genetic ancestry and an individual’s perceived ancestry can lead to confusion and a need to reevaluate one’s identity.^17,18^ However, individuals often prioritize their lived racial identity over reported genetic ancestry.^23,26^ Our findings highlight the importance of effectively communicating the current limitations of genetic ancestry testing to both patients and providers, particularly if it becomes more widely used in clinical care.

A significant concern regarding the increased use of genetic ancestry in medical care is the potential conflation of genetic ancestry as the sole explanation for health disparities among different groups.^3,4^ As research on the utility of genetic ancestry testing expands, it is crucial to consider other external factors that may contribute to differences in health risks and outcomes, such as social determinants of health (SDH).

The primary limitations of this study include a lack of diversity in the study population for the concordance and survey analyses, as most participants identified as White, and the small sample size of respondents from other ancestry groups, which may have affected our ability to detect statistically significant differences in responses. Additionally, the 3D Health participants were already enrolled in a research study, potentially biasing their responses in favor of genetic testing and its utility in the EHR.

Future studies on genetic ancestry testing should strive to include more diverse study populations to better identify inaccuracies in genetic ancestry reporting and determine best practices for calculating genetic ancestry. Moreover, further research is needed to investigate the potential utility of genetic ancestry data in clinical care algorithms and polygenic risk scores, which will help guide the integration of genetic ancestry into the EHR.

## Data Availability

All data produced in the present study are available upon reasonable request to the authors.

