## Supplemental Tables and Figure for "Integrating Genetic Ancestry into Clinical Care: Accuracy, Utility, and Stakeholder Views"

Supplemental Tables and Figures

**Supplemental Table 1. Genetic ancestry reporting from each genetic ancestry provider and Apex-reported race/ethnicity.**

| Genetic Ancestry Groupings | Ancestry 1 | Ancestry 2 | Apex-Reported Race/Ethnicity |
|---|---|---|---|
| African (AFR) | African Ancestry (SW US)<br>African Caribbean<br>Luhya (Kenya)<br>Mende (Sierra Leone)<br>Gambian Mendinka<br>Esan (S Nigeria)<br>Yoruba (Nigeria & Benin) | Not specified | Black or African American |
| Admixed American (AMR) | Puerto Rican<br>Peruvian<br>Colombian<br>Mexican | Not specified | Hispanic or Latino<br>American Indian or Alaska Native |
| East Asian (EAS) | Northern Chinese<br>Southern Chinese<br>Vietnamese<br>Dai (SW China)<br>Japanese | Not specified | Asian<br>Hawaii Native or Other Pacific Islander |
| European (EUR) | English & Scottish<br>Italian<br>Finnish<br>N&W European<br>Iberian (Spain & Portugal) | Not specified | White or Caucasian |
| South Asian (SAS) | Punjabi (Pakistan)<br>Bengali<br>Gujarati (W India)<br>Tamil Nadu-India<br>Telugu (SE India) | Not specified | Asian |

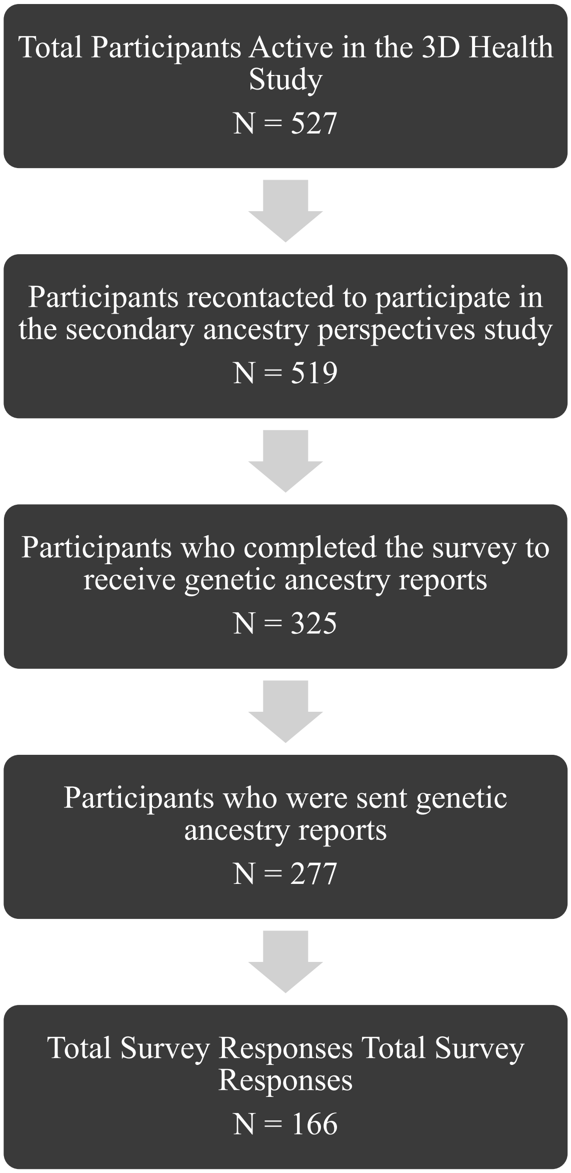


**Supplemental Figure 1. Flowchart of the 3D Health Study participants active and recontacted to receive genetic ancestry reports and complete the additional ancestry perspectives survey.**

**Supplemental Table 2. 3D Health Study Participant Demographics.**

| Variable | Completed Survey (N=166) n, % |
|---|---|
| Gender | |
| Man | 64 (38.6)% |
| Woman | 101 (60.8)% |
| Non-binary | 0 (0.0)% |
| Prefer not to answer | 1 (0.6)% |
| Age | |
| Below 30 | 7 (4.2)% |
| 30-45 | 49 (29.5)% |
| 45-60 | 27 (16.3)% |
| 60 or above | 82 (49.4)% |
| Prefer not to answer | 1 (0.6)% |
| Race/Ethnicity [a] | |
| American Indian/Alaska Native | 1 (0.6)% |
| Asian | 34 (20.5)% |
| Black/African American | 4 (2.4)% |
| Hispanic/Latino | 9 (5.4)% |
| Native Hawaiian/Pacific Islander | 0 (0.0)% |
| White | 124 (74.7)% |
| Other | 10 (6.0)% |
| Had Prior Genetic Ancestry testing | |
| Yes | 74 (44.6)% |
| No | 89 (53.6)% |
| Missing | 3 (1.8)% |

[a] Participants were allowed to select more than one race/ethnicity.

**Supplemental Table 3. UCSF Provider demographics**

| Variable | Completed Survey (N=31) n, % |
|---|---|
| Gender | |
| Man | 5 (16.1)% |
| Woman | 25 (80.6)% |
| Non-binary | 0 (0.0)% |
| Prefer not to answer | 1 (3.2)% |
| Age | |
| Below 30 | 0 (0.0)% |
| 30-45 | 18 (58.1)% |
| 45-60 | 11 (35.5)% |
| 60 or above | 2 (6.5)% |
| Prefer not to answer | 0 (0.0)% |
| Race/Ethnicity [a] | |
| American Indian/Alaska Native | 0 (0.0)% |
| Asian | 9 (29.0)% |
| Black/African American | 2 (6.5)% |
| Hispanic/Latino | 2 (6.5)% |
| Native Hawaiian/Pacific Islander | 0 (0.0)% |
| White | 21 (67.7)% |
| Other | 3 (9.7)% |
| Specialty | |
| Internal Medicine | 12 (38.7)% |
| Family Medicine | 2 (6.5)% |
| Genetics | 1 (3.2)% |
| Genetic Counseling | 14 (45.2)% |
| Other | 2 (6.5)% |
| Had Prior Genetic Ancestry testing | |
| Yes | 5 (16.1)% |
| No | 26 (83.9)% |

[a] Participants were allowed to select more than one race/ethnicity.

**Supplemental Table 4. Concordance between genetic ancestry calculations.** Genetic ancestry results from two commercial genetic testing providers and two 3rd-party analyses were compared using genetic samples in the UCSF 3D Health Study. The agreement was determined by the % of overlapping ancestry from each ancestry location (European, African, Asian, and Admixed American). P values were determined using a Wilcox Rank sum test.

| Ancestry 1 vs. Ancestry 3 (n=460) | | | | | | | |
|---|---|---|---|---|---|---|---|
| Agreement | Min | Median | Max | Mean | | P-values | |
| Overall | 0.00% | 54.65% | 94.91% | 57.00% | | | |
| Genetic Ancestry | | | | | Afr | As | Adm |
| European (333) | 26.00% | 53.41% | 78.03% | 53.34% | 6.0e-5** | 6.6e-22** | 0.63 |
| African (9) | 53.57% | 79.99% | 94.91% | 76.18% | | 0.37 | 9.8e-4** |
| Asian (75) | 36.45% | 78.26% | 89.94% | 72.90% | | | 5.7e-6** |
| Admixed American (18) | 0.00% | 53.34% | 72.78% | 52.52% | | | |
| Ancestry 1 vs. Ancestry 4 (n=460) | | | | | | | |
| Agreement | Min | Median | Max | Mean | | | |
| Overall | 0.00% | 54.79% | 95.21% | 57.26% | | | |
| Genetic Ancestry | | | | | | | |
| European (333) | 32.27% | 52.96% | 79.67% | 53.12% | 2.5e-3** | 1.6e-29** | 1.3e-3** |
| African (9) | 47.92% | 79.99% | 95.21% | 73.69% | | 0.75 | 0.076 |
| Asian (75) | 38.66% | 78.67% | 89.94% | 74.53% | | | 9.4e-6** |
| Admixed American (18) | 0.00% | 62.70% | 74.77% | 58.21% | | | |
| Ancestry 2 vs. Ancestry 3 (n=465) | | | | | | | |
| Agreement | Min | Median | Max | Mean | | | |
| Overall | 33.50% | 99.87% | 100.00% | 95.80% | | | |
| Genetic Ancestry | | | | | | | |
| European (337) | 33.50% | 99.90% | 100.00% | 97.24% | 7.5e-4** | 4.0e-3** | 1.3e-9** |
| African (9) | 92.73% | 94.53% | 97.10% | 94.97% | | 2.1e-4** | 6.0e-4** |
| Asian (74) | 78.60% | 100.00% | 100.00% | 97.62% | | | 3.7e-10** |
| Admixed American (18) | 38.72% | 65.15% | 100.00% | 66.83% | | | |
| Ancestry 2 vs. Ancestry 4 (n=465) | | | | | | | |
| Agreement | Min | Median | Max | Mean | | | |
| Overall | 58.68% | 100.00% | 100.00% | 95.70% | | | |
| Genetic Ancestry (n) | | | | | | | |
| European (337) | 58.68% | 100.00% | 100.00% | 97.33% | 3.5e-5** | 2.4e-15** | 2.3e-12** |
| African (9) | 83.13% | 95.52% | 98.62% | 94.37% | | 0.040* | 3.1e-3** |
| Asian (74) | 71.67% | 97.97% | 100.00% | 94.65% | | | 6.3e-6** |
| Admixed American (18) | 58.72% | 75.65% | 100.00% | 78.32% | | | |
| Ancestry 3 vs. Ancestry 4 (n=474) | | | | | | | |
| Agreement | Min | Median | Max | Mean | | | |
| Overall | 58.77% | 99.41% | 100.00% | 97.05% | | | |
| Genetic Ancestry (n) | | | | | | | |

| | | | | | | | |
|---|---|---|---|---|---|---|---|
| European (344) | 71.90% | 99.72% | 100.00% | 98.76% | 2.3e-4** | 1.3e-13** | 6.0e-9** |
| African (9) | 81.30% | 98.48% | 99.01% | 96.39% | | 0.90 | 6.2e-3* |
| Asian (76) | 62.90% | 97.91% | 100.00% | 93.54% | | | 1.7e-3** |
| Admixed American (18) | 58.77% | 89.40% | 100.00% | 86.08% | | | |

*P < 0.05; **P<0.005

**Supplemental Table 5. Pare-wise comparisons of the concordance of the four ancestries when compared to Apex-reported ancestry.** McNemar's test was used on each pairwise comparison.

| | Threshold | | |
|---|---|---|---|
| Comparison | 50% | 75% | 90% |
| Ancestry 1/2 | 5.0e-24** | 1.7e-70** | 9.6e-74** |
| Ancestry 1/3 | 1.4e-21** | 6.4e-71** | 3.8e-81** |
| Ancestry 1/4 | 1.5e-22** | 1.4e-71** | 3.1e-82** |
| Ancestry 2/3 | 0.21 | >0.99 | 4.0e-7** |
| Ancestry 2/4 | 0.23 | 0.45 | 1.1e-8** |
| Ancestry 3/4 | >0.99 | 0.131 | 0.041* |

*P < 0.05; **P<0.005

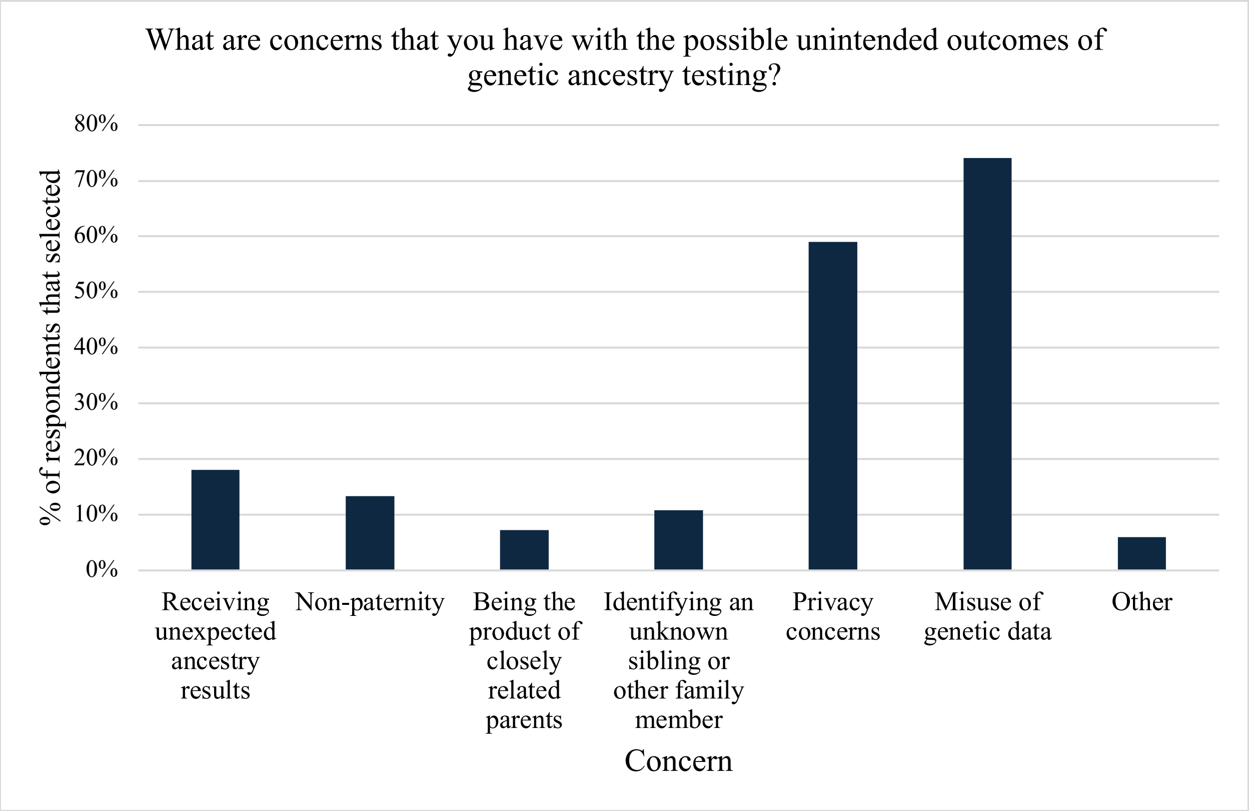


**Supplemental Figure 2. 3D Health Participant concerns about the possible unintended outcomes of genetic ancestry testing.**

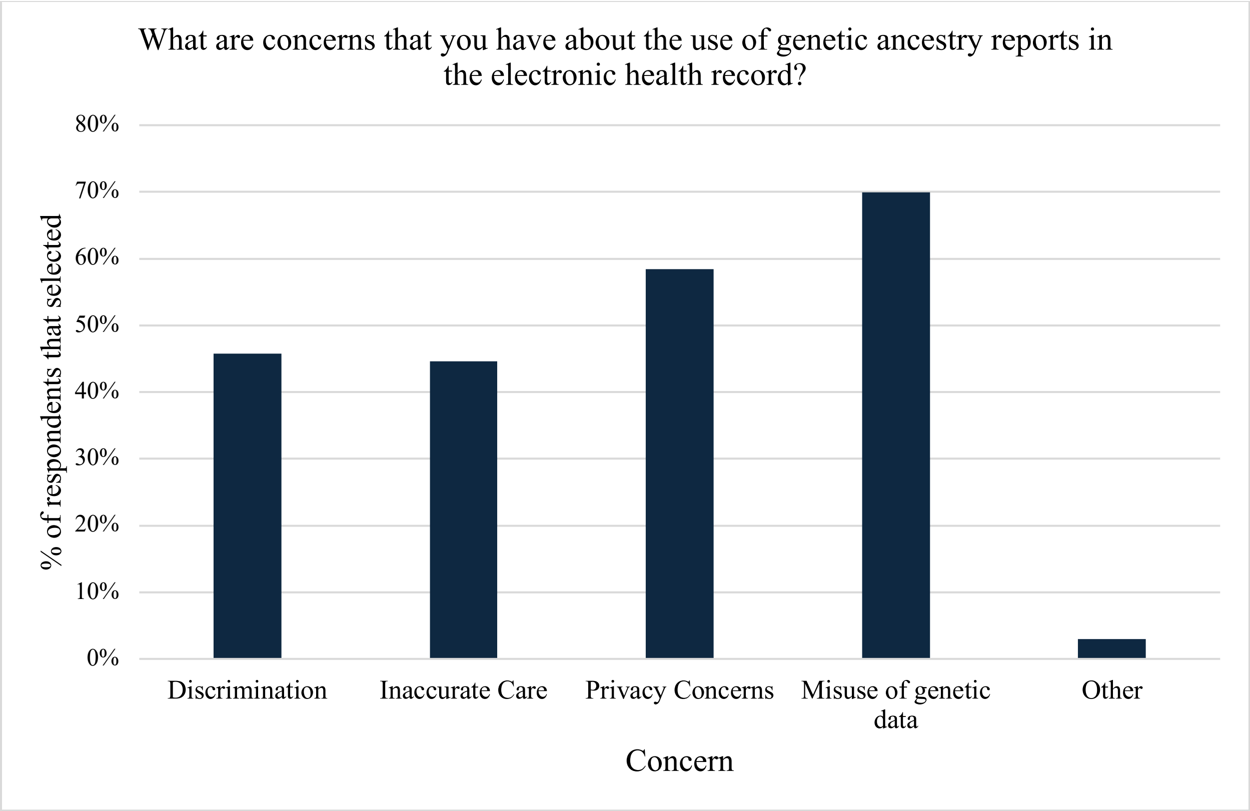


**Figure 3. 3D Health Participant concerns on the use of genetic ancestry reports in the electronic health record.**

**Supplemental Table 6. Provider perspectives on the integration of genetic ancestry into the electronic health record.** Responses were totaled for the overall responses from all providers and then calculated based on the participant's self-identified race/ethnicity. Providers who selected more than one race/ethnicity that did not fit into one of the general ancestry categories were placed into other. A Fisher's Exact test was used to determine if there was a statistically significant difference in responses between ancestry groups.

| Genetic ancestry reports should be in the electronic health record. | Strongly disagree | Somewhat disagree | Neutral | Somewhat agree | Strongly agree | P-value |
|---|---|---|---|---|---|---|
| | 2 (6.5)% | 6 (19.4)% | 16 (51.6)% | 7 (22.6)% | 0 (0.0)% | |
| European (n=18) | 1 (5.6)% | 4 (22.2)% | 8 (44.4)% | 5 (27.8)% | 0 (0.0)% | |
| African (n=1) | 0 (0.0)% | 0 (0.0)% | 1 (100.0)% | 0 (0.0)% | 0 (0.0)% | |
| Asian (n=7) | 1 (14.3)% | 1 (14.3)% | 4 (57.1)% | 1 (14.3)% | 0 (0.0)% | 0.9913 |
| Admixed American (n=1) | 0 (0.0)% | 0 (0.0)% | 1 (100.0)% | 0 (0.0)% | 0 (0.0)% | |
| Other (n=4) | 0 (0.0)% | 1 (25.0)% | 2 (50.0)% | 1 (25.0)% | 0 (0.0)% | |
