## Supplemental Text for "Integrating Genetic Ancestry into Clinical Care: Accuracy, Utility, and Stakeholder Views"

**Supplemental Text 1. 3D Health Participant Survey**

Demographics

Which gender do you identify as?

a. Man

b. Woman

c. Non-binary

d. Prefer not to answer

e. Other (please specify): ________________

What is your age?

a. Below 30

b. 30-45

c. 45-60

d. 60 or above

e. Prefer not to answer

Please specify your racial/ethnic identity. (You may choose more than one)

a. American Indian/Alaskan Native

b. Asian

c. Black/African American

d. Hispanic/Latino

e. Native Hawaiian/Other Pacific Islander

f. White

g. Other (please specify): ____________

Have you had genetic ancestry testing done prior to participating in the UCSF 3D Health Study?

a. Yes

i. What company did you use?

- [Open text field]

ii. Would you be willing to share your results with our study?

- Yes
- No

iii. What prompted you to undergo genetic ancestry testing? (You may choose more than one)

- Personal curiosity
- To gain more insights about myself
- To discover more about my family history
- For health-related reasons
- Other (please specify): __________

b. No

i. What is the reason you haven't undergone genetic ancestry testing? (You may choose more than one)

- Lack of interest
- High cost
- Concerns with the accuracy of the test
- Worries over privacy
- Fear of discrimination
- Lack of knowledge about genetic ancestry testing
- Other (please specify): __________

Impact of Genetic Ancestry Results

Were your ancestry test results surprising or unexpected?

a. Yes

b. No

c. Somewhat

Were your ancestry test results undesired or distressing?

a. Yes

b. No

c. Somewhat

Do your ancestry test results change your perceptions of your cultural roots?

a. Yes

b. No

c. Somewhat

Do your ancestry test results change the likelihood that you would travel to certain parts of the world in the future?

a. Yes

b. No

c. Somewhat

Do your ancestry test results change how you view certain cultures or world regions?

a. Yes

b. No

c. Somewhat

Would you say your ancestry test results have reshaped your personal identity?

a. Yes

b. No

c. Somewhat

Do you plan to share your ancestry test results with your family members?

a. Yes

b. No

c. Maybe

Will you provide or discuss your ancestry test results with your physician or a healthcare provider?

a. Yes

b. No

c. Maybe

If you had genetic ancestry testing previously, do you perceive your current ancestry test results to be different than your previous results? (If this is your first genetic ancestry test, please skip this question.)

a. Yes

b. No

c. Somewhat

d. Not applicable

Does the experience of undergoing genetic ancestry testing make you more or less likely to have other genetic tests in the future?

a. More Likely

b. Less Likely

c. No Change

What are the advantages to learning your estimated genetic ancestry?

a. [Open text field]

What are the disadvantages to learning your estimated genetic ancestry?

a. [Open text field]

<u>Genetic Ancestry Testing and Medical Care</u>

Reflect on your own genetic ancestry reports and this statement; genetic ancestry reports are known to vary because of how they are calculated.

For the following questions, please rate your agreement on a scale from 1 (strongly disagree) to 5 (strongly agree)

a. Genetic ancestry reports are accurate.
b. Genetic ancestry reports will allow for more personalized care.
c. Genetic ancestry reports should be used by healthcare providers to make health related decisions.
d. Genetic ancestry reports should be used by healthcare providers to better understand their patients as a whole.
e. Genetic ancestry reports should be in the electronic health record.

If genetic ancestry is included in the electronic health record, how would you want it to be integrated?

a. Hidden and only used for assessing risks that take ancestry into account
b. Visible to all providers
c. Not sure
d. Other: ______________

Concerns of Genetic Ancestry Testing

What are concerns that you have with the possible unintended outcomes of genetic ancestry testing? (Select all that apply)

a. Receiving unexpected ancestry results

b. Non-paternity

c. Being the product of closely related parents

d. Identifying an unknown sibling or other family member

e. Privacy concerns

f. Misuse of genetic data

g. Other (please specify): ________

What are concerns that you have about the use of genetic ancestry reports in the electronic health record? (Select all that apply)

a. Discrimination

b. Inaccurate care

c. Privacy concerns

d. Misuse of genetic data

e. Other (please specify): _________

Do you expect that genetic ancestry reports will be used to discriminate against certain groups of people?

a. Yes

b. No

Additional Comments:

a. [Open text field]

**Supplemental Text 2. UCSF Provider Survey**

Demographics

Which gender do you identify as?

f. Man

g. Woman

h. Non-binary

i. Prefer not to answer

j. Other (please specify): ________________

What is your age?

f. Below 30

g. 30-45

h. 45-60

i. 60 or above

j. Prefer not to answer

Please specify your racial/ethnic identity. (You may choose more than one)

h. American Indian/Alaskan Native

i. Asian

j. Black/African American

k. Hispanic/Latino

l. Native Hawaiian/Other Pacific Islander

m. White

n. Other (please specify): ____________

Which medical specialty do you practice?

a. Internal Medicine

b. Family Medicine

c. Genetics

d. Genetic counseling

e. Other (please specify): ____________

Have you ever undergone genetic ancestry testing?

a. Yes

i. Which service provider did you use?

- [Open text field]

ii. What prompted you to undergo genetic ancestry testing? (You may choose more than one)

- Personal curiosity
- To gain more insights about myself
- To discover more about my family history
- For health-related reasons
- Other (please specify): __________

b. No

i. What's the reason you haven't undergone genetic ancestry testing? (You may choose more than one)

- Lack of interest
- High cost
- Doubts about the test's accuracy
- Worries over privacy
- Fear of discrimination

- Other (please specify): __________

Impact of Genetic Ancestry Results on Medicine

Reflect on your knowledge regarding the influence of race and genetic ancestry on healthcare for the following questions.

(For the following questions, please rate your agreement on a scale from 1 (strongly disagree) to 5 (strongly agree))

a. Race influences health.
b. Race, to the extent that it corresponds with genetic ancestry, influences health.
c. Genetic variations between races explain health disparities.
d. Racial categorizing is important for biomedical research.
e. Genetics professionals should be knowledgeable about the correlation between race, genetics, and health.
f. Health professionals should comprehend key concepts about relationship between race, genetics, and health.
g. Development of medications targeted towards specific racial groups is a necessary advancement towards personalized medicine.
h. Race should be a consideration in the diagnosis of certain conditions or diseases.
i. Genetic ancestry should be a factor in the diagnosis of certain conditions or diseases.
j. Race should be considered in the treatment of certain conditions or diseases.
k. Genetic ancestry should be a factor in the treatment of certain conditions or diseases.

Thoughts on Genetic Ancestry Reports in Electronic Health Records

For the following question, please rate your agreement on a scale from 1 (strongly disagree) to 5 (strongly agree)

a. Genetic ancestry reports should be in the electronic health record.

If genetic ancestry is included in the electronic health record, how would you want it to be integrated?

a. Hidden, but used for risk assessment that incorporates ancestry

b. Visible to all healthcare providers

c. Not sure

d. Other (please specify): ______________

Implications of Genetic Ancestry Testing on Your Clinical Practice

For the following questions, please rate your agreement on a scale from 1 (strongly disagree) to 5 (strongly agree)

a. I feel capable of accurately interpreting genetic ancestry data and understanding its health implications.

b. I feel confident in explaining genetic ancestry results to patients.

How frequently do you utilize genetic ancestry testing results in your practice?

a. Never

b. Occasionally

c. Sometimes

d. Most of the time

e. Always

In what areas of patient care do you use genetic ancestry test results?

a. Clinical care algorithms

b. Evaluating the risk of monogenic conditions (e.g., Sickle Cell Trait/Disease, Tay Sachs)

c. Evaluating the risk polygenic or multifactorial conditions (e.g., Diabetes, Cancer, Heart Disease)
d. Interpreting genetic testing results
e. Other (please specify): _______________
f. I do not apply genetic ancestry testing in my practice.

How frequently do you anticipate using genetic ancestry testing results in the future?

a. Never
b. Occasionally
c. Sometimes
d. Most of the time
e. Always

In what areas of patient care would you utilize genetic ancestry test results in the future?

a. Clinical care algorithms
b. Evaluating the risk of monogenic conditions (e.g., Sickle Cell Trait/Disease, Tay Sachs)
c. Evaluating the risk of polygenic or multifactorial conditions (e.g., Diabetes, Cancer, Heart Disease)
d. Interpreting genetic testing results
e. Other (please specify): _______________
f. I do not foresee myself using genetic ancestry testing in my practice.

Concerns with Genetic Ancestry Testing

What concerns do you have regarding the potential unexpected outcomes of genetic ancestry testing? (You may choose more than one)

a. Unforeseen ancestry results

b. Non-paternity

c. Being the product of closely related parents

d. Identification of an unknown sibling or other family member

e. Privacy concerns

f. Misuse of genetic data

g. Other (please specify): ________

What are concerns that you have about the use of your genetic ancestry reports in the electronic health record? (Select all that apply)

a. Discrimination

b. Compromised care quality

c. Privacy concerns

d. Misuse of genetic data

e. Other (please specify): _________

Do you believe that genetic ancestry reports could be exploited to discriminate against certain groups of people?

a. Yes

b. No

Additional comments:

a. [Open text field]
